# Detecting Fifth Metatarsal Fractures on Radiographs through the Lens of Smartphones: A FIXUS AI Algorithm

**DOI:** 10.1101/2025.07.18.25331772

**Authors:** Atta Taseh, Aaditya Shah, Mani Eftekhari, Alexandra Flaherty, Alireza Ebrahimi, Sumner Jones, Varun Nukala, Ara Nazarian, Gregory Waryasz, Soheil Ashkani-Esfahani

## Abstract

**Background:** Fifth metatarsal (5MT) fractures are common but challenging to diagnose, particularly with limited expertise or subtle fractures. Deep learning shows promise but faces limitations due to image quality requirements. This study develops a deep learning model to detect 5MT fractures from smartphone-captured radiograph images, enhancing accessibility of diagnostic tools.

**Methods:** A retrospective study included patients aged >18 with 5MT fractures (n=1240) and controls (n=1224). Radiographs (AP, oblique, lateral) from Electronic Health Records (EHR) were obtained and photographed using a smartphone, creating a new dataset (SP). Models using ResNet 152V2 were trained on EHR, SP, and combined datasets, then evaluated on a separate smartphone test dataset (SP-test).

**Results:** On validation, the SP model achieved optimal performance (AUROC: 0.99). On the SP-test dataset, the EHR model’s performance decreased (AUROC: 0.83), whereas SP and combined models maintained high performance (AUROC: 0.99).

**Conclusions:** Smartphone-specific deep learning models effectively detect 5MT fractures, suggesting their practical utility in resource-limited settings.

## 1 Introduction

Metatarsal fractures are among the most common foot fractures, with an incidence rate of 75.4 per 100,000 people per year. Fifth metatarsal (5MT) fractures account for more than 80% of single metatarsal fractures.^1^ These fractures can be categorized into three zones: Zone 1 occurs at the proximal tubercle and is typically treated non-operatively, whereas Zone 2 (Jones fractures) and Zone 3 (proximal diaphyseal fractures) require more detailed assessment for appropriate clinical decision-making.^2^ The specific anatomy of the arteries supplying the 5MT bone makes it prone to bone union complications, necessitating meticulous diagnosis and treatment.^3^ Diagnosis is based on physical examination and plain radiographs - anteroposterior (AP), lateral, and oblique views - which confirm the fracture location and type. ^4,5,6,7,8^

The delay in the treatment process gives rise to complications like malunion and increases patients’ chances of long-term morbidity.^9,10^ This is a frequent scenario for malpractice claims against clinicians, which can happen in high-pressure settings such as emergency departments as well as resource-limited (RL) settings.^11,12^ While seeking second opinions through teleradiology has been effective in reducing error rates, the infrastructural demands of implementing a Picture Archiving and Communication System (PACS) present significant challenges in RL settings.^13^ The literature highlights key issues, including unstable power and network connections, limited image storage capacity, and insufficient knowledge of information technology in these medical settings.^14^ Smartphone communication has emerged as a practical alternative for obtaining second opinions, proving to be promising despite the reduced quality of radiological images.^15^ However, the shortage of radiologists in RL places remains a critical issue.^13^ Given the advances in artificial intelligence (AI) and fracture detection, smartphone cameras could become a valuable tool in aiding clinical decision-making, improving accessibility to expert input, and facilitating faster data transfer and communication. ^16,17,18^

In this study, we hypothesize that a deep learning AI algorithm trained on smartphone images of radiographs will achieve superior performance in detecting 5MT fractures compared to conventionally trained models on high-quality images. Validating this hypothesis through rigorous testing would establish a foundation for potentially leveraging AI technology to enhance diagnostic capabilities in RL or high-pressure settings.

## 2 Materials and Methods

### 2.1 Study Design and Population

After obtaining approval from the Institutional Review Board (IRB #), a retrospective case-control study was conducted at a tertiary institution, including data from three hospitals. Due to the retrospective nature of the study, informed consent was waived by the IRB.

International Classification of Diseases-10 [S92.301 (A, B), S92.302 (A, B), S92.309 (A, B), S92.351 (A, B), S92.352 (A, B), S92. 353 (A, B), S92.354 (A, B), S92.355 (A, B), S92.356 (A, B)] to query the data from the institution’s data repository. Patients aged ≥18 years with an isolated 5MT fracture were included in the case (Fx) group, and those aged ≥18 years without a fracture or severe deformity in the foot area were included in the control (NoF) group (Table 1). Severe deformities such as hallux valgus, hammer toes, metatarsus adductus, and Charcot disease were excluded from the NoF group.

**Table 1.** Eligibility criteria for study groups.

| Group | Inclusion Criteria | Exclusion Criteria |
| --- | --- | --- |
| Case | - Age ≥ 18 years<br>- Isolated 5<sup>th</sup> metatarsal fracture<br>- Available standard X-rays views* | - Age <18 years<br>- Presence of fractures in multiple regions of the foot<br>- Presence of hardware, cast, or artifact<br>- Presence of bone cysts, or lesions<br>- Healing fracture (>2 weeks)<br>- Insufficient X-ray views |
| Control | - Age ≥ 18 years<br>- No fracture in the foot area<br>- Available standard X-rays views* | - Age <18 years<br>- Presence of fracture in the foot<br>- Severe deformity, degenerative changes, or soft tissue injuries<br>- Presence of hardware, cast, or artifact<br>- Presence of bone cysts, or lesions<br>- Insufficient X-ray views |

\*Anteroposterior, Oblique, and Lateral

### 2.2 Data Collection and Preparation

Three experienced orthopedic researchers reviewed the dataset for inclusion/exclusion criteria. Radiographs, along with CT scans, MRIs where available, and clinical radiological notes, were carefully examined to confirm diagnoses. Once the eligibility was confirmed, radiographs were extracted from the institution’s Electronic Health Records (EHR) system and saved in Portable Network Graphic (PNG) format. Smartphone photography was used to capture radiograph images, employing different Android and iOS smartphones and screens to ensure variability, which was also considered as a manual augmentation technique. All photographs were taken in a dark room, with screens at maximum brightness and the cellphone positioned approximately 20 centimeters from the screen.

For the Fx set, we created an EHR dataset consisting of three-view foot radiographs (AP, lateral, and oblique) from 1240 patients. Two versions of the EHR dataset were created by photographing radiographs displayed on computer screens using smartphone cameras, one set with an Android device and the other with an iOS device. (Figure 1). Five percent of each version was reserved for final testing, ensuring no duplicates for smartphone test (SP-test). The SP-test images were also removed from the original EHR dataset. A smartphone dataset (SP) was then created by combining 50% of the remaining images from each version, without duplicate patients. Lastly, 50% of the SP and EHR datasets were mixed to build the combined dataset, avoiding duplicates. The same process was followed for the NoF group, including a dataset of 1224 healthy individuals.

**Figure 1.**
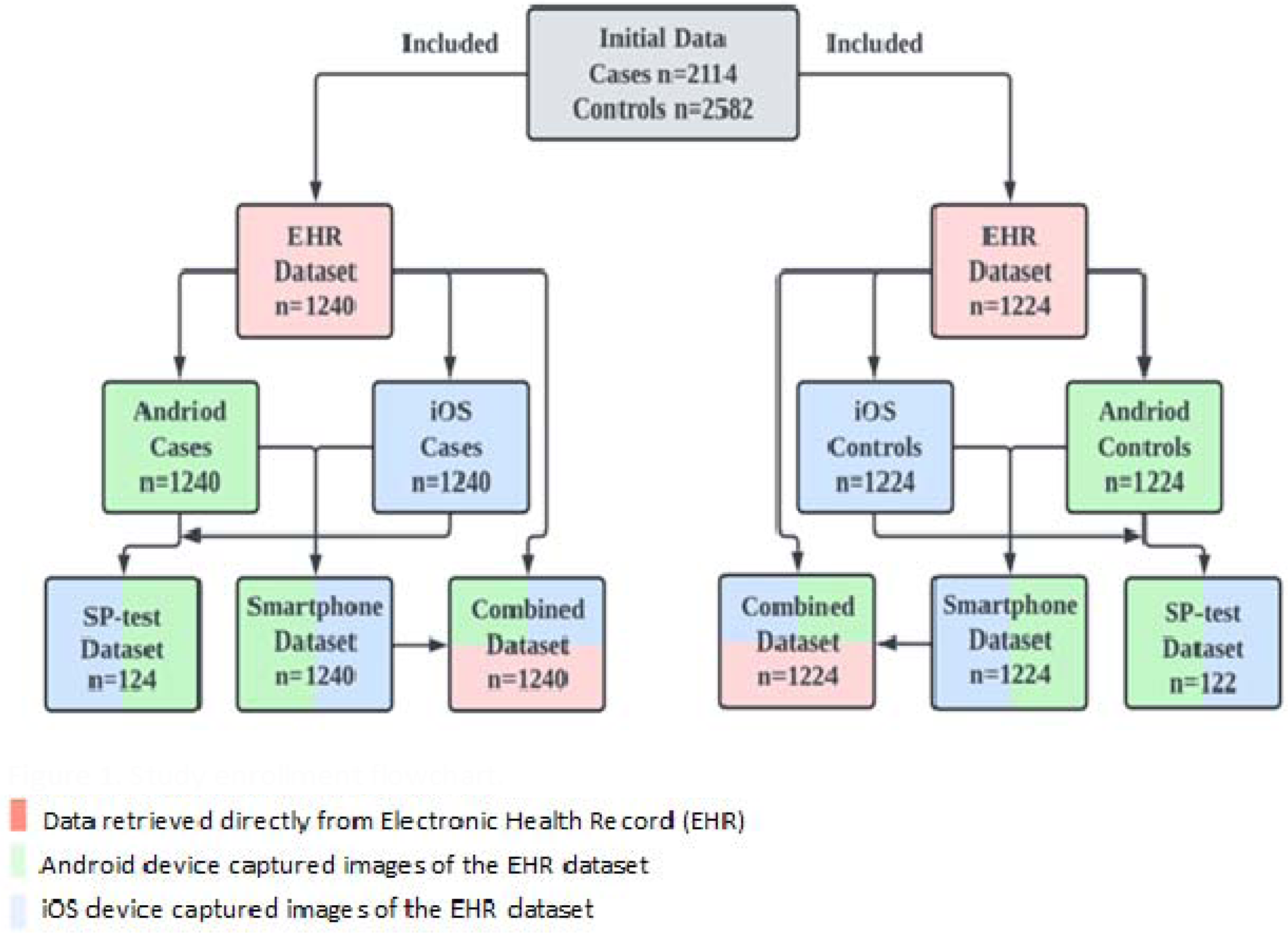
Study enrollment flowchart. Note: Color should be used for Figure 1.

In the preprocessing phase, the images were resized to a uniform dimension of 600×600 pixels with the 3 color channels, red, green, and blue, aligning with common practices in medical image processing where consistent image dimensions are essential for model input.^19^ Normalization of pixel values to the range [0,1] was preformed to standardize the data, a step that is important in deep learning (DL) to ensure model convergence and performance optimization.^20^

### 2.3 Model Development

Three convolutional neural networks (CNNs) were designed for the classification task using the Keras library and TensorFlow platform. The ResNet 152-v2 architecture was used for the base of the models, with an additional dense layer with 1024 filters (with ReLU activation), a dropout layer with 0.4 dropout rate, and a final dense layer with sigmoid activation added for the binary classification.^21^ The models were compiled using the Adam optimizer with an initial learning rate of 1e-4, with binary cross-entropy as the loss function.^22^ After training for fifty epochs, the last 20 layers of the ResNet model would be unfrozen, and the model was retrained with a learning rate of 1e-6. Additional layers were trained if the model training accuracy was not improving.^23^ The models were trained using a 90:10 training and validation split on their respective datasets and then tested on SP-test dataset. This allowed for comprehensive training and performance evaluation. The combined model is publicly available for testing at the following link: <u>FIXUS-AI</u>

### 2.4 Statistical Analysis

Data analysis was conducted using SPSS (Version 28.0, IBM, USA). We assessed the normality of continuous data using the Kolmogorov-Smirnov and Shapiro-Wilk tests. Depending on the distribution of the data, the Mann-Whitney U test was applied for comparisons. Results are presented as median (Interquartile Range, IQR), and percentages (%). The performance of the models was evaluated based on key metrics such as the Area Under the Receiver Operating Characteristics Curve (AUROC), sensitivity, specificity, positive predictive value (PPV), negative predictive value (NPV), and Youden’s J. index A p-value of 0.05 was considered statistically significant.

## 3 Results

A total of 1240 records with a median age of 56 (36, 68) years and 1224 with a median age of 62 (51, 72) years were included in the Fx and NoF groups (p<0.001). Although most participants in both groups were white, there was a significant difference in racial composition of the study groups (p<0.001; Table 2).

**Table 2.** Demographic characteristics of the study groups. Data is presented as median (IQR) or percentage.

| Characteristic |  | Cases | Controls | P-value |
| --- | --- | --- | --- | --- |
| Age (years) |  | 56 (36, 68) | 62 (51, 72) | <0.001<sup>a</sup> |
| Gender (Female) |  | 75.5% | 72.1% | 0.174<sup>b</sup> |
| Race | White | 84.8% | 92.2% | <0.001<sup>b</sup> |
|  | Non-white | 15.2% | 7.8% |  |

<sup>a</sup> Mann-Whitney U test
<sup>b</sup> Chi Square test

Our models were evaluated for the detection of 5MT fractures on X-rays in various views. All three models showed high performance when evaluated on their validation datasets, with the SP model achieving the best performance with an AUROC of 0.99 and a Youden’s J of 0.95 (Table 3; Figure 2). On the SP-test dataset, the SP and combined models maintained high performance, while the EHR model showed a decline, with an AUROC of 0.83 and a Youden’s J of 0.88 (Table 3; Figure 2).

**Figure 2.**
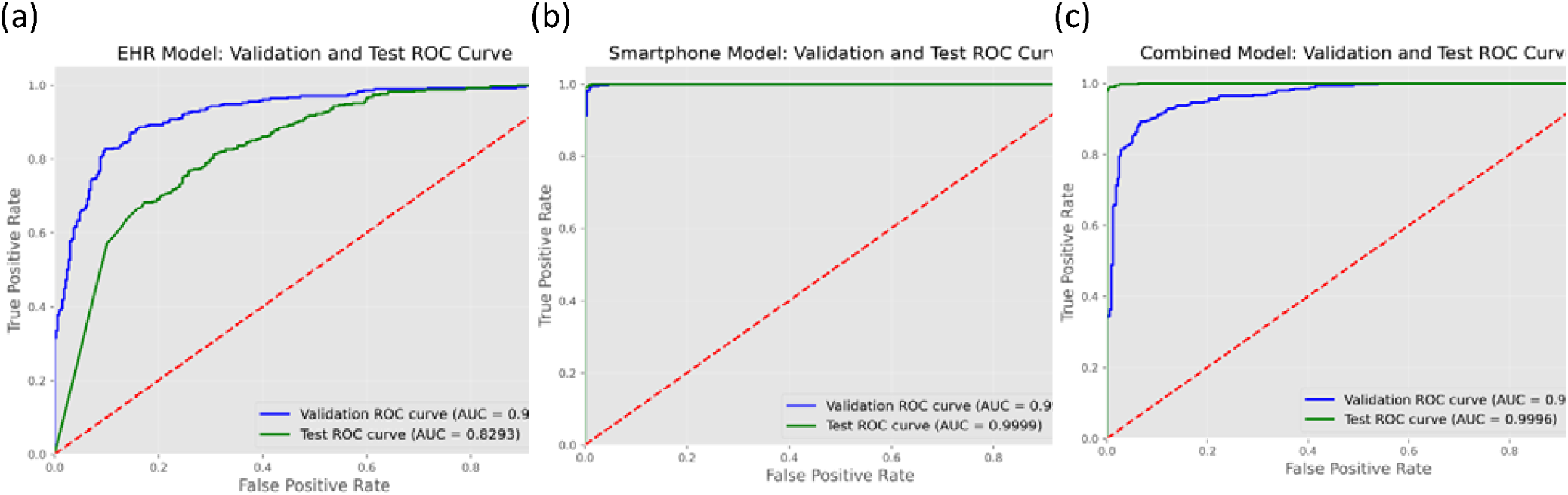
Comparison of receiver operating characteristic (ROC) curves for study models, evaluated on both validation and smartphone datasets (SP-test). (a) Electronic Health Record (EHR) model developed on data directly retrieved from HER system; (b) Smartphone model, developed on smartphone-captured images of EHR data; (c) Combined model developed using both EHR and smartphone datasets Note: Color should be used for Figure 2.

**Table 3.**
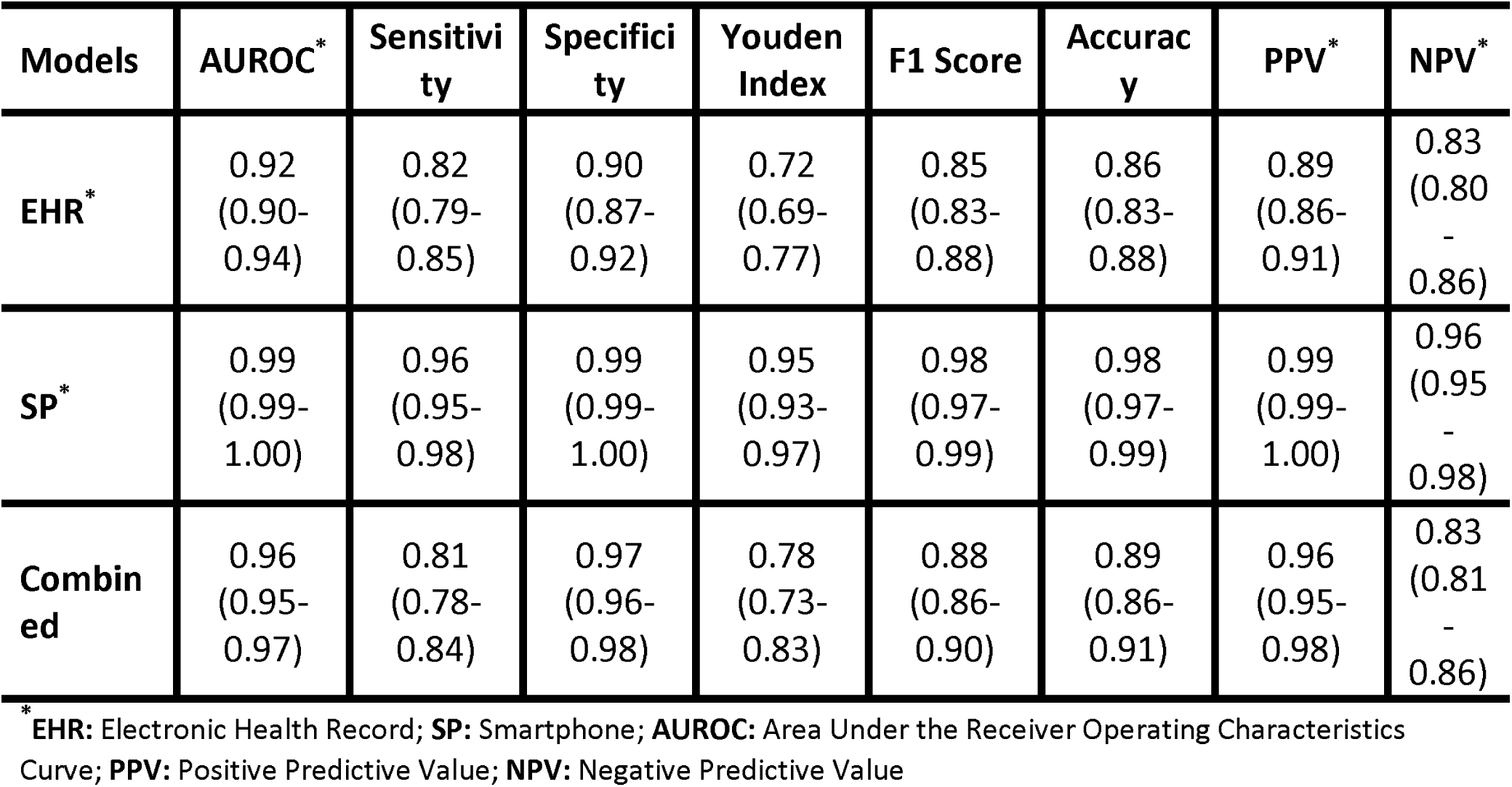
Performance metrics of study models tested on original validation datasets, with 95% confidence intervals shown in parentheses.

| Models | AUROC* | Sensitivity | Specificity | Youden Index | F1 Score | Accuracy | PPV* | NPV* |
| --- | --- | --- | --- | --- | --- | --- | --- | --- |
| **EHR\*** | 0.92<br>(0.90-0.94) | 0.82<br>(0.79-0.85) | 0.90<br>(0.87-0.92) | 0.72<br>(0.69-0.77) | 0.85<br>(0.83-0.88) | 0.86<br>(0.83-0.88) | 0.89<br>(0.86-0.91) | 0.83<br>(0.80-0.86) |
| **SP\*** | 0.99<br>(0.99-1.00) | 0.96<br>(0.95-0.98) | 0.99<br>(0.99-1.00) | 0.95<br>(0.93-0.97) | 0.98<br>(0.97-0.99) | 0.98<br>(0.97-0.99) | 0.99<br>(0.99-1.00) | 0.96<br>(0.95-0.98) |
| **Combined** | 0.96<br>(0.95-0.97) | 0.81<br>(0.78-0.84) | 0.97<br>(0.96-0.98) | 0.78<br>(0.73-0.83) | 0.88<br>(0.86-0.90) | 0.89<br>(0.86-0.91) | 0.96<br>(0.95-0.98) | 0.83<br>(0.81-0.86) |

\* **EHR:** Electronic Health Record; **SP:** Smartphone; **AUROC:** Area Under the Receiver Operating Characteristics Curve; **PPV:** Positive Predictive Value; **NPV:** Negative Predictive Value

**Table 4.** Performance metrics of study models evaluated on an independent smartphone-captured dataset (SP-test) not used during model development, with 95% confidence intervals shown in parentheses.

| Model<br>s | AUROC<sup>*</sup> | Sensitivity | Specificity | Youden<br>Index | F1<br>Score | Accuracy | PPV<sup>*</sup> | NPV<sup>*</sup> |
| --- | --- | --- | --- | --- | --- | --- | --- | --- |
| **EHR<sup>\*</sup>** | 0.83<br>(0.80-0.85) | 0.98<br>(0.97-0.99) | 0.90<br>(0.87-0.92) | 0.88<br>(0.85-0.91) | 0.85<br>(0.83-0.88) | 0.86<br>(0.83-0.88) | 0.89<br>(0.86-0.91) | 0.83<br>(0.80-0.86) |
| **SP<sup>\*</sup>** | 0.99<br>(0.99-1.00) | 0.98<br>(0.98-0.99) | 0.99<br>(0.99-1.00) | 0.98<br>(0.97-0.99) | 0.99<br>(0.99-1.00) | 0.99<br>(0.99-1.00) | 0.99<br>(0.99-1.00) | 0.98<br>(0.98-0.99) |
| **Combined** | 0.99<br>(0.99-1.00) | 0.97<br>(0.96-0.98) | 0.99<br>(0.99-1.00) | 0.96<br>(0.94-0.98) | 0.98<br>(0.98-0.99) | 0.98<br>(0.98-0.99) | 0.99<br>(0.99-1.00) | 0.97<br>(0.96-0.98) |

<sup>\*</sup> **EHR:** Electronic Health Record; **SP:** Smartphone; **AUROC:** Area Under the Receiver Operating Characteristics Curve; **PPV:** Positive Predictive Value; **NPV:** Negative Predictive Value

## 4 Discussion

Plain radiographs are the first-line diagnostic tool for detecting various orthopaedic injuries. Despite their widespread use, interpretation challenges often arise, particularly in RL settings, or when handled by inexperienced clinicians or care providers. This study assessed the diagnostic accuracy of AI algorithms for detecting 5MT fractures on smartphone-captured images of radiographs, developed using combinations of datasets. Our findings showed that AI models trained exclusively on smartphone images achieved superior diagnostic performance, highlighting the potential of tailored training for smartphone-based diagnostic tools.

The application of AI in orthopedics, especially for fracture detection, is a well-studied area that has shown performance comparable to human raters. Vertebral, hip, and upper extremity fractures are among the most extensively researched areas.^24^ The majority of these studies utilized plain radiographs as the modality of interest, resulting in a pooled sensitivity of 94% and a specificity of 92% for fracture detection.^25^ However, the use of deep learning for fracture detection in the foot area remains comparatively underexplored.^26^ The complex anatomy of the foot and the subtle presentation of certain fractures pose unique challenges. Nevertheless, emerging research has begun to address this gap. Kim et al. evaluated the efficiency of different convolutional neural networks for foot fracture detection and reported an AUROC of up to 0.95 using ensemble methods.^27^ Their study, however, included a relatively small sample of 436 single-view images combining different fractures in the foot area. Similarly, Wang et al. conducted a study using 1,151 lower extremity radiographs to investigate the application of deep learning in detecting stress fractures and grading their severity.^28^ They reported an overall AUROC of 0.94, missing only 9.7% of lower grade fractures, compared to 39.9% missed by human raters. However, none of these studies provided a subgroup analysis of each specific fracture in the foot area, likely due to a lack of sufficient sample size. In our study, we addressed this limitation by specifically training models to detect 5MT fractures using a larger sample size. Our models achieved an AUROC of up to 0.99, sensitivity of 0.96, and specificity of 0.99 confirming the results of previous studies and underscoring the potential of deep learning algorithms in accurately detecting specific foot fractures.

With over three billion users worldwide, smartphones are among the most accessible technologies today.^29^ Given the increasing importance of diagnosing, incorporating it into smartphones is an effective way to enhance their utility, accessibility, and improve healthcare quality. For instance, Yang et al. developed an algorithm using 1,819 smartphone-acquired images of thick blood smears to detect malaria, achieving an AUROC of 98.39%.^30^ More relevant to our study, Rangarajan et al. developed a smartphone application to diagnose COVID-19 using images of chest radiographs.^31^ Their best-performing model reported a positive likelihood ratio of 27.3 when evaluated on smartphone-based images of radiographs comprising 271 healthy, 441 pneumonia, and 48 COVID-19 cases. However, due to the small size of their dataset, factors such as variations in camera quality, screen quality, and user technique pose significant risks to reliable performance outside controlled environments. Furthermore, they employed the publicly available datasets and techniques like data augmentation and Generative Adversarial Networks (GANs) to synthesize new data and address the issue of sample size limitation for their training dataset. While these techniques are invaluable, they may lead to overfitting or generate unrealistic image features that do not generalize well to real-world scenarios, especially in clinical diagnosis.^32^ In our study, we aimed to address these issues by including a larger sample size and creating smartphone datasets using various brands of cellphones, screens, and users to replicate real-world scenarios. Our results demonstrated higher performance of the SP and combined models when tested using smartphone images, confirming the value of training models with smartphone-acquired images for diagnosing diseases using image data when developing smart device applications.

This study had several notable strengths, including a larger sample size compared to similar studies in the field and the creation of SP datasets under diverse conditions, enhancing the model’s applicability to real-world scenarios. However, some limitations should be noted. The generalizability of the findings may be somewhat restricted, as data were primarily collected from three hospitals in the same geographic region, which may limit applicability to other populations or clinical settings with different demographics and imaging equipment.

## 5 Conclusions

Deep learning models demonstrate superior performance in detecting 5MT from smartphone-captured images when trained on specific datasets. Our results suggest that developing models tailored for smart device-based imaging could offer a practical diagnostic solution in medical settings with limited access to specialized expertise or advanced imaging equipment. Nonetheless, further prospective validation is necessary to confirm these models’ utility and robustness in clinical applications.

## Data Availability

All data produced in the present study are available upon reasonable request to the authors and approval from the IRB.

## Acknowledgements

We would like to acknowledge and express our sincere gratitude to all the patients whose clinical data provided the foundation for this research.

## References

1. Herterich V, Hofmann L, Böcker W, Polzer H, Baumbach SF. Acute, isolated fractures of the metatarsal bones: an epidemiologic study. Arch Orthop Trauma Surg. 2022;143(4):1939–1945. doi:10.1007/s00402-022-04396-3

2. Chloros GD, Kakos CD, Tastsidis IK, Giannoudis VP, Panteli M, Giannoudis PV. Fifth metatarsal fractures: an update on management, complications, and outcomes. EFORT Open Rev. 2022;7(1):13–25. doi:10.1530/EOR-21-0025

3. Smidt KP, Massey P. 5th Metatarsal Fracture. In: StatPearls. StatPearls Publishing; 2025. Accessed April 22, 2025. http://www.ncbi.nlm.nih.gov/books/NBK544369/

4. Guly HR. Diagnostic errors in an accident and emergency department. Emerg Med J. 2001;18(4):263–269. doi:10.1136/emj.18.4.263

5. Kung JW, Melenevsky Y, Hochman MG, et al. On-Call Musculoskeletal Radiographs: Discrepancy Rates Between Radiology Residents and Musculoskeletal Radiologists. Am J Roentgenol. 2013;200(4):856–859. doi:10.2214/AJR.12.9100

6. Donald JJ, Barnard SA. Common patterns in 558 diagnostic radiology errors. J Med Imaging Radiat Oncol. 2012;56(2):173–178. doi:10.1111/j.1754-9485.2012.02348.x

7. Fitschen-Oestern S, Lippross S, Lefering R, et al. Missed foot fractures in multiple trauma patients. BMC Musculoskelet Disord. 2019;20(1):121. doi:10.1186/s12891-019-2501-8

8. Jibri Z, Mukherjee K, Kamath S, Mansour R. Frequently Missed Findings in Acute Ankle Injury. Semin Musculoskelet Radiol. 2013;17(04):416–428. doi:10.1055/s-0033-1356471

9. Miele V, Galluzzo M, Trinci M. Missed Fractures in the Emergency Department. In: Romano L, Pinto A, eds. Errors in Radiology. Springer Milan; 2012:39–50. doi:10.1007/978-88-470-2339-0_5

10. Hallas P, Ellingsen T. Errors in fracture diagnoses in the emergency department – characteristics of patients and diurnal variation. BMC Emerg Med. 2006;6(1):4. doi:10.1186/1471-227X-6-4

11. Whang JS, Baker SR, Patel R, Luk L, Castro A. The Causes of Medical Malpractice Suits against Radiologists in the United States. Radiology. 2013;266(2):548–554. doi:10.1148/radiol.12111119

12. Berlin L. Defending the “Missed” Radiographic Diagnosis. Am J Roentgenol. 2001;176(2):317–322. doi:10.2214/ajr.176.2.1760317

13. Schoeman R, Haines M. Radiologists’ experiences and perceptions regarding the use of teleradiology in South Africa. South Afr J Radiol. 2023;27(1). doi:10.4102/sajr.v27i1.2647

14. Abbas R, Singh Y. PACS Implementation Challenges in a Public Healthcare Institution: A South African Vendor Perspective. Healthc Inform Res. 2019;25(4):324. doi:10.4258/hir.2019.25.4.324

15. Ntja U, Janse Van Rensburg J, Joubert G. Diagnostic accuracy and reliability of smartphone captured radiologic images communicated via WhatsApp®. Afr J Emerg Med. 2022;12(1):67–70. doi:10.1016/j.afjem.2021.11.001

16. Meena T, Roy S. Bone Fracture Detection Using Deep Supervised Learning from Radiological Images: A Paradigm Shift. Diagnostics. 2022;12(10):2420. doi:10.3390/diagnostics12102420

17. Kalmet PHS, Sanduleanu S, Primakov S, et al. Deep learning in fracture detection: a narrative review. Acta Orthop. 2020;91(2):215–220. doi:10.1080/17453674.2019.1711323

18. Su Z, Adam A, Nasrudin MF, Ayob M, Punganan G. Skeletal Fracture Detection with Deep Learning: A Comprehensive Review. Diagnostics. 2023;13(20):3245. doi:10.3390/diagnostics13203245

19. University of Craiova, Romania, Ivanescu RC. A statistical evaluation of the preprocessing medical images impact on a deep learning network’s performance. Ann Univ Craiova -Math Comput Sci Ser. 2022;49(2):411–421. doi:10.52846/ami.v49i2.1641

20. Sane P, Agrawal R. Pixel normalization from numeric data as input to neural networks: For machine learning and image processing. In: 2017 International Conference on Wireless Communications, Signal Processing and Networking (WiSPNET). IEEE; 2017:2221–2225. doi:10.1109/WiSPNET.2017.8300154

21. He K, Zhang X, Ren S, Sun J. Deep Residual Learning for Image Recognition. Published online 2015. doi:10.48550/ARXIV.1512.03385

22. Kingma DP, Ba J. Adam: A Method for Stochastic Optimization. Published online 2014. doi:10.48550/ARXIV.1412.6980

23. Xiao X, Bamunu Mudiyanselage T, Ji C, Hu J, Pan Y. Fast Deep Learning Training through Intelligently Freezing Layers. In: 2019 International Conference on Internet of Things (iThings) and IEEE Green Computing and Communications (GreenCom) and IEEE Cyber, Physical and Social Computing (CPSCom) and IEEE Smart Data (SmartData). IEEE; 2019:1225–1232. doi:10.1109/iThings/GreenCom/CPSCom/SmartData.2019.00205

24. Bhatnagar A, Kekatpure AL, Velagala VR, Kekatpure A. A Review on the Use of Artificial Intelligence in Fracture Detection. Cureus. Published online April 16, 2024. doi:10.7759/cureus.58364

25. Jung J, Dai J, Liu B, Wu Q. Artificial intelligence in fracture detection with different image modalities and data types: A systematic review and meta-analysis. Frasch MG, ed. PLOS Digit Health. 2024;3(1):e0000438. doi:10.1371/journal.pdig.0000438

26. Gupta P, Kingston KA, O’Malley M, Williams RJ, Ramkumar PN. Advancements in Artificial Intelligence for Foot and Ankle Surgery: A Systematic Review. Foot Ankle Orthop. 2023;8(1):24730114221151079. doi:10.1177/24730114221151079

27. Kim T, Goh TS, Lee JS, Lee JH, Kim H, Jung ID. Transfer learning-based ensemble convolutional neural network for accelerated diagnosis of foot fractures. Phys Eng Sci Med. 2023;46(1):265–277. doi:10.1007/s13246-023-01215-w

28. Wang Y, Li Y, Lin G, et al. Lower-extremity fatigue fracture detection and grading based on deep learning models of radiographs. Eur Radiol. 2022;33(1):555–565. doi:10.1007/s00330-022-08950-w

29. Susanto AP, Winarto H, Fahira A, et al. Building an artificial intelligence-powered medical image recognition smartphone application: What medical practitioners need to know. Inform Med Unlocked. 2022;32:101017. doi:10.1016/j.imu.2022.101017

30. Yang F, Poostchi M, Yu H, et al. Deep Learning for Smartphone-Based Malaria Parasite Detection in Thick Blood Smears. IEEE J Biomed Health Inform. 2020;24(5):1427–1438. doi:10.1109/JBHI.2019.2939121

31. Rangarajan AK, Ramachandran HK. A preliminary analysis of AI based smartphone application for diagnosis of COVID-19 using chest X-ray images. Expert Syst Appl. 2021;183:115401. doi:10.1016/j.eswa.2021.115401

32. Kim J, Park H. Limited Discriminator GAN using explainable AI model for overfitting problem. ICT Express. 2023;9(2):241–246. doi:10.1016/j.icte.2021.12.014

